# Longitudinal Evaluation of mRNA Vaccinated Subjects using a Quidel Multianalyte Point-of-Care SARS-CoV-2 IgG Immunoassay

**DOI:** 10.1101/2021.05.06.21256544

**Authors:** Xi Chen, Sarika Agarwal, Stewart Hoelscher, Richard Egan, Dipesh Jaiswal, Adonis Stassinopoulos, Robert Reed, Jason McClure, Werner Kroll

**Author notes:** Corresponding Author: Robert Reed, Research and Development, 9975 Summers Ridge Road, San Diego, CA 92121.

## Abstract

Infection from SARS-CoV-2 elicits an immune response to the nucleocapsid (N) and spike proteins (subunits S1 and S2). In this study, we set out to understand the utility of the multiplexed Quidel Sofia 2 SARS-CoV-2 IgG Antibody Fluorescent Immuno-Assay (FIA) that measures IgG antibodies against these three primary SARS-CoV-2 antigens from a single sample in 15 minutes. Using this assay with samples that were collected prior to the COVID-19 pandemic (n=816) and diseased state samples (n=99), the specificities for the three antigens were 98.4-99.9% and 98.0-100.0%, respectively. A longitudinal study was designed to collect weekly fingerstick, venous whole blood, serum and plasma samples from subjects vaccinated with the Moderna or Pfizer/BioNtech mRNA vaccines. The majority of these enrolled subjects had no known prior infection while a subset was known to have had prior COVID-19 infection. We found that the fingerstick whole blood samples performed as effectively as serum, plasma, and venous whole blood samples with a 95.8-99.5% agreement allowing physicians in a near-patient setting to rapidly provide results to their patients. Additionally, as this assay measures an IgG response against three viral proteins, S1, S2 and N, we were able to characterize immune response between *i)* naturally infected subjects, *ii*) vaccinated subjects with no prior infection, *iii*) vaccinated subjects with known prior infection, and *iv*) vaccinated subjects with prior asymptomatic exposure/infection. The Quidel Sofia 2 SARS-CoV-2 IgG FIA will aid in providing insights to the protective humoral responses as an increasing number of the world population is vaccinated against SARS-CoV-2.

## Introduction

As the 2019 Coronavirus (COVID-19) spread rapidly across the world to millions of persons, the administration of a safe and effective vaccine to counter this pandemic became critical. Because of the faster development cycles and the ease of production of mRNA vaccines, efforts focused on developing RNA/molecular-based vaccine delivery systems to target SARS-CoV-2. According to [CDC](https://covid.cdc.gov/covid-data-tracker/#vaccinations), by the middle of April 2021, over 190 million Moderna and Pfizer-BioNTech doses had been administered in the United States. The two vaccines released have similar designs. Both are mRNA vaccines encoding full length modified Spike (S) protein using lipid nanoparticle (LNP)-formulation as a delivery vehicle (1-3). The S protein is a key surface protein on the SARS-CoV-2 surface and the primary target for neutralizing antibodies (4). The injected packaged mRNA/LNP, produces a modified S protein that was shown to be highly immunogenic (5-7). Both vaccines produce cellular as well as humoral immune responses thus inhibiting virus binding to the angiotensin-converting enzyme 2 (ACE-2) receptor (8, 9). Published reports show that both vaccines have a protective efficacy of 95% two weeks after receiving the second dose (3, 10, 11). As required by the FDA, the mRNA vaccines follow a two-dose regimen, a 2x 100 µg dose with Moderna and a 2x 30 µg dose with Pfizer-BioNTech (3, 12-14).

The Moderna and Pfizer-BioNTech vaccines specifically target the pre-fusion S protein structure by developing modified forms that ensure the S protein does not change its conformation helping to elicit a humoral response to the pre-fusion form of the S protein (1, 2, 8, 15). The S protein consists of two subunits: S1 and S2. The receptor binding domain (RBD) region is part of the S1 subunit. The interaction between RBD and the human ACE-2 receptor is required for viral entry into the host cell. The S2 subunit plays a role in the entry into a host cell but lacks significant neutralizing epitopes. The nucleocapsid (N) protein is highly immunogenic and is an abundantly expressed protein during infection (16). High levels of IgG antibodies against N have been detected in sera from COVID-19 patients (16). SARS-CoV-2 serology tests have been developed that target the immune response to different combinations of the S (RBD, S1 and S2 subunits) and N proteins. While most commercial assays target IgG, some also detect IgM and IgA. Assay types include visually read rapid tests and semi-quantitative ELISA assays, and are used with different venous sample matrices (17-20). As more information on the immune response to specific viral proteins becomes available, it may become possible to distinguish between vaccinated and non-vaccinated subjects with and without prior exposure.

With the availability of two mRNA vaccines, each expressing a modified full-length S protein, the focus of a new multiplex serology assay is to independently report the IgG immune response to each of the three, S1, S2 and N viral antigens. The primary objective of this study was to better understand vaccination-induced IgG antibody response using Quidel’s multiplex lateral flow, Sofia 2 SARS-CoV-2 IgG antibody fluorescent immunoassay (FIA). Further, the assay can distinguish between subjects with prior exposure to the virus pre-vaccination and from subjects without prior exposure pre-vaccination. Additionally, the test identified individuals that had asymptomatic infections or exposures prior to vaccination resulting in a more robust immune responses upon vaccination. As the vaccines do not code for the N protein, reactivity to the N protein can only be derived from a COVID-19 infection rather than vaccination. An additional advantage of this assay is the use of a fingerstick sample as opposed to a venous sample as a convenient and effective collection method.

## Results and Discussion

The rollout of mRNA vaccines in the US has implications for the interpretation of SARS-CoV-2 serology testing. Prior to the introduction of vaccines, serology tests that detect antibodies specific for major viral proteins would indicate exposure to SARS-CoV-2. After vaccination with the two mRNA vaccines, individuals were routinely monitored for seropositivity against the S protein. This study provides insights into the immune responses elicited from subjects that were vaccinated with either the Moderna or Pfizer-BioNTech vaccine that either had no prior exposure or had prior exposure to SARS-CoV-2.

### Reactivity and Specificity of the assay

The Sofia 2 SARS-CoV-2 IgG antibody FIA was evaluated using serum samples from ten RT-PCR confirmed SARS-CoV-2 infected subjects. A longitudinal antibody response against S1, S2 and N is shown in Figure 1. Within 8-12 days, a strong IgG response was observed against all three antigens from serum samples from these infected subjects. Specificity of Sofia 2 SARS-CoV-2 IgG antibody FIA was confirmed using 816 COVID-19 negative serum and plasma samples that were collected and stored frozen prior to the COVID-19 pandemic. The observed specificity was determined to be 98.4% (803/816) for N, 99.9% (815/816) for S1 and 99.0% (808/816) for S2 (Table 1). In addition, a panel of 99 donors from non-COVID-19 diseased state serum and plasma samples that included subjects characterized with exposure to seasonal coronavirus were also evaluated (Table 1). The specificity of the diseased state samples was 98.0% (97/99) for N, 100.0% (99/99) for S1 and 99.0% (98/99) for S2.

**Table 1.** Specificity of S1, S2 and N with Pre-COVID-19 Samples and *Disease State Samples.

| Samples | Antigens | Samples | Negative | Positive | Specificity |
| --- | --- | --- | --- | --- | --- |
| Pre-COVID 19 Negative | N | 816 | 803 | 13 | 98.4% |
| Pre-COVID 19 Negative | S1 | 816 | 815 | 1 | 99.9% |
| Pre-COVID 19 Negative | S2 | 816 | 808 | 8 | 99.0% |
| *Disease State Samples | N | 99 | 97 | 2 | 98.0% |
| *Disease State Samples | S1 | 99 | 99 | 0 | 100.0% |
| *Disease State Samples | S2 | 99 | 98 | 1 | 99.0% |

\* Serum/Plasma samples from subjects with known infections other than SARS-CoV-2.

**Figure 1.**
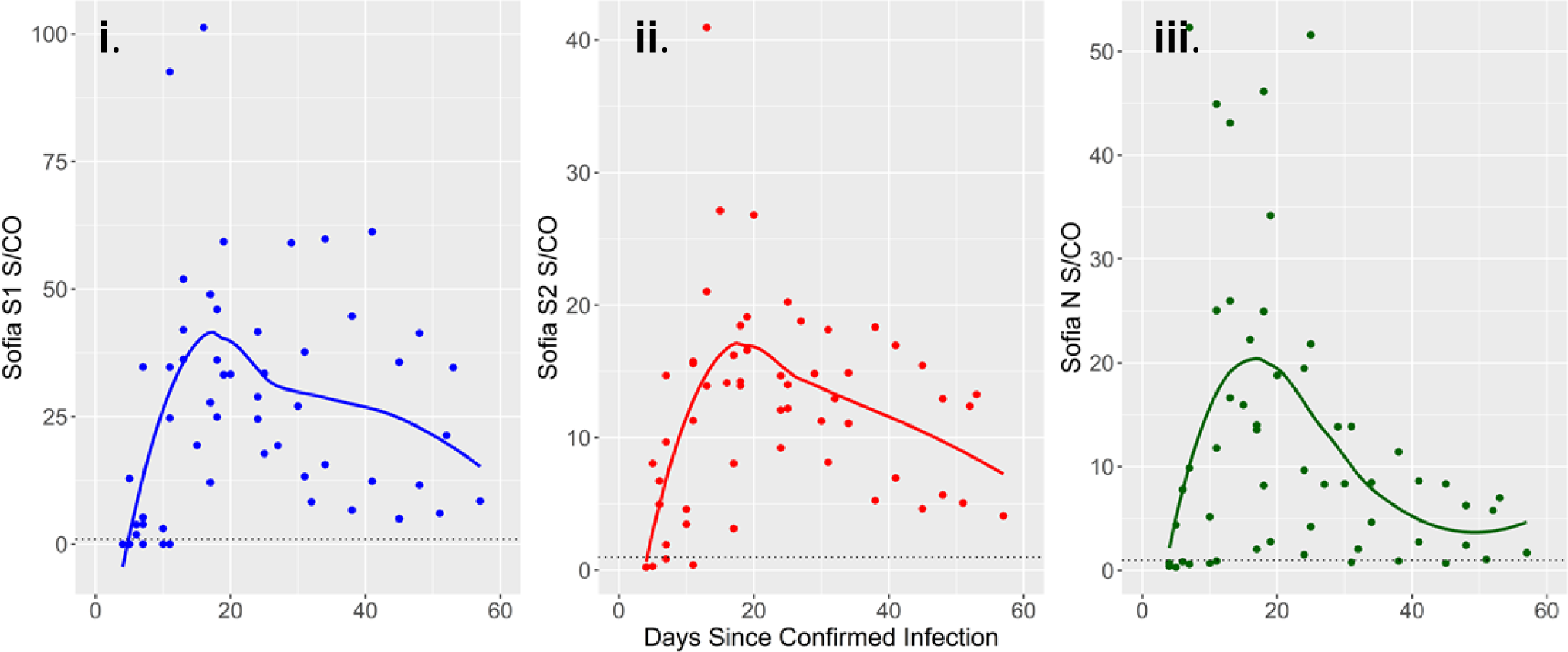
Longitudinal profile of IgG response against SARS-CoV-2 S1, S2 and N antigens from subjects with RT-PCR confirmed COVID-19 diagnosis with no vaccination. Panels i, ii, and iii represent IgG levels against S1 (i), S2 (ii), and N (iii) from banked serum samples from confirmed COVID-19 infected subjects (n=10). IgG response against S1 is represented by a solid blue line, S2 by a solid red line and N by a green solid line. LOESS fit is used to smooth each line (21). Each blue, red and green dot represents an IgG response against S1, S2 and N. The y-axis represents the S/CO values for S1, S2 and N. S/CO numbers above 1, marked by the horizontal dashed black line, represent a positive IgG response against S1, S2 and N. Please note the different scales on the y-axis. The x-axis represents the time elapsed in days since COVID-19 infection confirmation.

### Enrollment and Collection of samples

A multi-center, IRB approved study was designed to collect samples prior to and after the first and second Moderna and Pfizer-BioNTech vaccine doses. Subjects with prior infection, or no known infection prior to the first vaccination were included in this study. Subjects included in the study were male or female, age 18 years or older, and must have had the first blood draw within 7 days of receiving the first COVID-19 mRNA vaccine. Subjects were also required to obtain the second vaccination according to the manufacturer’s requirement. Following consent, demographics, symptoms, and health history were collected from each subject (Table 2). Matched fingerstick, venous whole blood, plasma, and serum samples were collected from each subject on a weekly basis. Samples were collected for 3 weeks after the second vaccination dose.

**Table 2.**
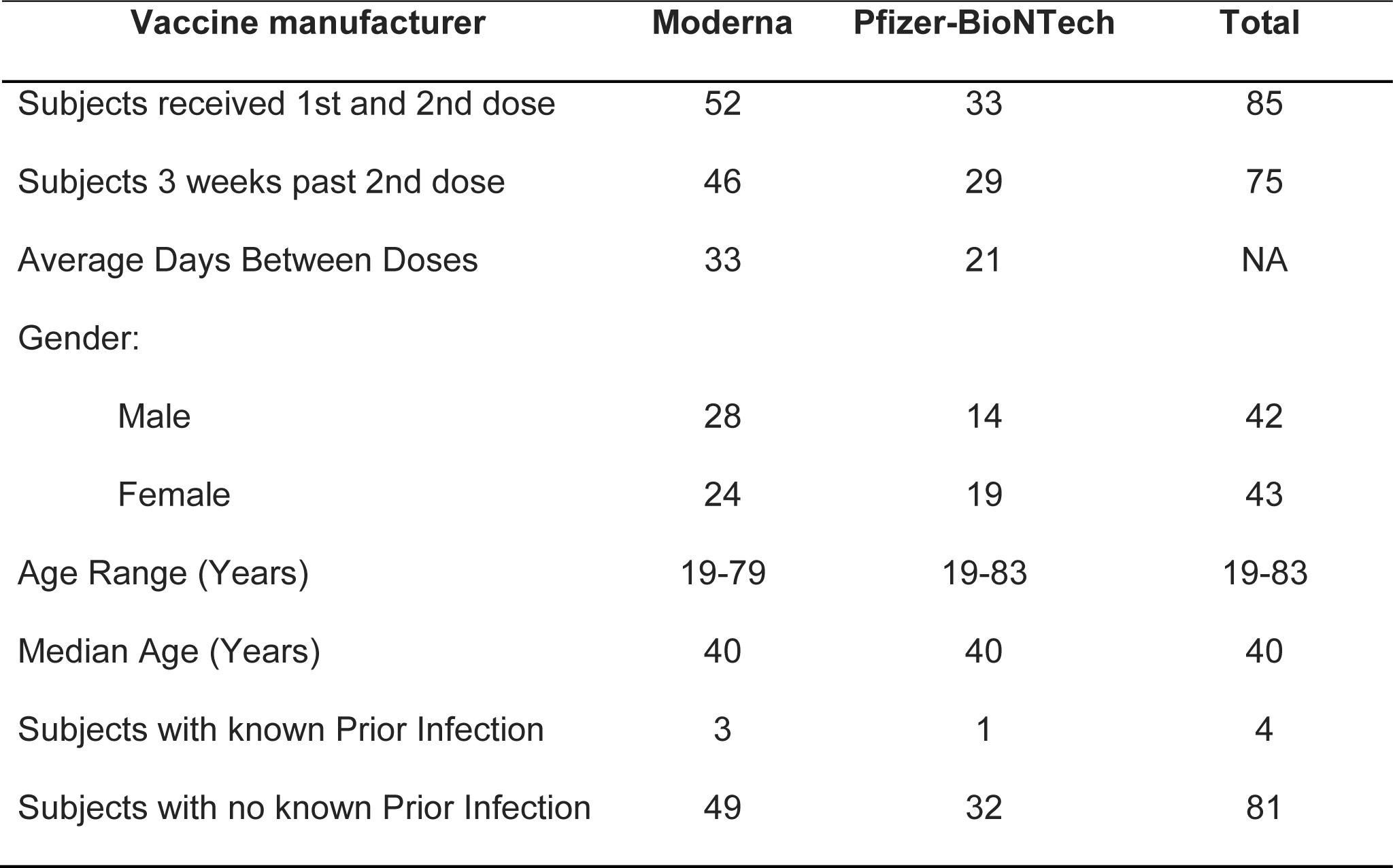
Demographics of the study participants.

Of a total of 85 enrolled subjects, 46 subjects that received the Moderna vaccine and 29 subjects that received the Pfizer-BioNTech vaccine were 3 weeks past the second dose. All four samples types from each subject were tested in parallel on the day of collection using the Quidel Sofia 2 SARS-CoV-2 IgG Antibody FIA.

### Matrix Comparison

The Sofia 2 SARS-CoV-2 IgG Antibody FIA is designed to be used with fingerstick whole blood, venous whole blood, serum, or plasma samples. In point-of-care settings, the fingerstick whole blood sample is advantageous due to a simple and short collection process that requires a 25µl whole blood sample collected using a disposable plastic capillary tube. In this study we compared the IgG response against S1, S2 and N antigens from each of the four sample types over time. Sample matrix equivalency was demonstrated by qualitative percent agreement of the results from fingerstick, plasma, serum, and venous whole blood samples. The qualitative agreement of the fingerstick whole blood results was greater than 95% with all combinations of sample matrices for all analytes as shown in Table 3. Based on the robust sample equivalency results, the fingerstick sample data was used for analysis of the longitudinal study.

**Table 3.** Percent agreement between fingerstick whole blood samples and plasma, serum, and venous whole blood samples.

| Sample Type | % Agreement |  |  |
| --- | --- | --- | --- |
|  | S1 – Fingerstick | S2 – Fingerstick | N – Fingerstick |
| Plasma | 98.5% (588/597) | 96.8% (578/597) | 96.3% (575/597) |
| Serum | 98.7% (586/594) | 96.1% (571/594) | 95.8% (569/594) |
| Venous Whole blood | 99.5% (594/597) | 97.0% (579/597) | 97.0% (579/597) |

### Longitudinal monitoring of antibody response for vaccinated subjects

The IgG immune response was measured against S1, S2 and N from subjects vaccinated with Moderna (n=30) and Pfizer-BioNTech (n=24) but with no known prior exposure to SARS-CoV-2. The primary IgG immune response was against the S1 antigen (Figure 2A). IgG antibody response was not observed against the S2 and N proteins (Figure 2B and C). We observed that a majority of subjects vaccinated using either mRNA vaccine had an immune response to S1 2-3 weeks after the first vaccine dose. With time points from both vaccine groups normalized to the second vaccine dose, the average antibody response to S1 was similar approximately 2 weeks after the second dose.

**Figure 2.**
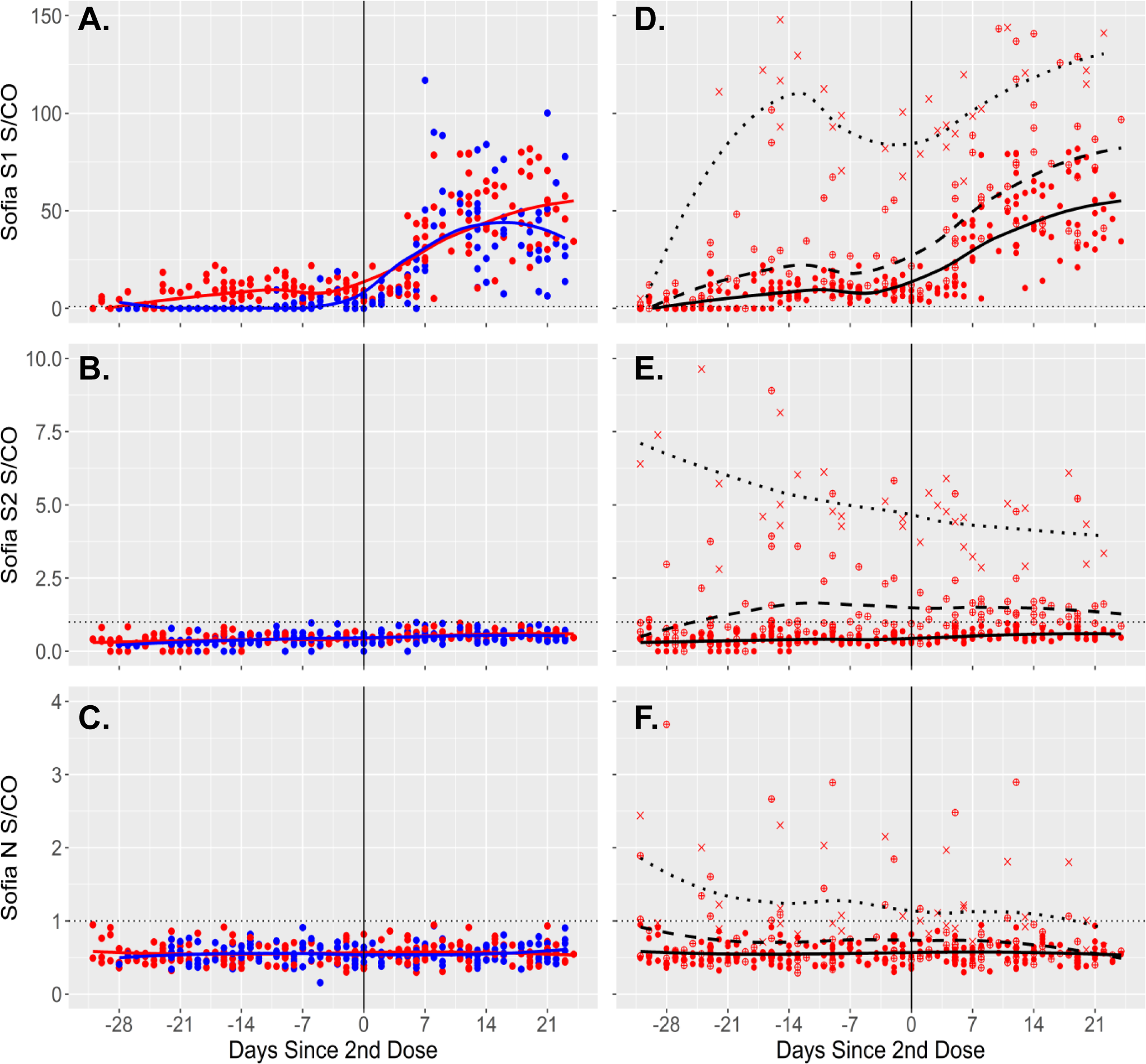
Longitudinal IgG response from fingerstick samples against SARS-CoV-2 S1, S2 and N antigens after vaccination with mRNA vaccines. Panels A, B, and C represent IgG levels against S1 (A), S2 (B), and N (C) from subjects immunized with Moderna (Red solid line) and Pfizer-BioNTech (Blue solid line) vaccines with no prior COVID-19 infection. Each red dot represents a Moderna vaccinated sample, and each blue dot represents a Pfizer-BioNTech vaccinated sample. Panels D, E, and F represent IgG levels against S1 (D), S2 (E), and N (F). Samples with no known prior COVID-19 infection are represented by a solid black line, samples with known prior COVID-19 infection are reflected by a bold dotted black line and samples with possible prior exposure to the virus are shown by a dashed black line. A red dot, a red cross and a red circled cross represents a non-infected, a prior-infected and a possible prior exposed sample. LOESS fit is used to smooth each line (21). The solid vertical black line shows the day the second vaccine dose was administered. S/CO numbers above 1, marked by the horizontal dashed black line, represent a positive IgG response against S1, S2 and N.

In comparison, prior exposure to SARS-CoV-2 should elicit an antibody response against S1, S2 and N proteins. In this study, the immune response for subjects previously exposed to SARS-CoV-2 and vaccinated with Moderna (n=3) and Pfizer-BioNTech (n=1) were higher (Figure 2D) than the IgG immune response from subjects with no prior exposure to the virus. Following the first vaccination, S1 IgG response of the pre-exposed group was rapid and visibly higher (S/CO > 100) than the non-exposed subject response (S/CO ∼15). The increased S1 IgG levels in the pre-exposed group was observed 4-5 days after the first vaccination compared to the non-exposed subjects at 13 days. Following the second dose, the pre-exposed subjects S1 response increased to a similar high response seen after the first vaccination dose (Figure 2D). A [previous publication](https://www.eurosurveillance.org/content/10.2807/1560-7917.ES.2021.26.6.2100096) similarly noted a strong reaction to the first COVID-19 vaccine dose in subjects with prior COVID-19 infection. Previously infected subjects in this study had detectable antibodies against S2 and N antigens as well (Figure 2E and F).

As the S2 and N levels did not increase or spike after the first or the second vaccine dose, they are likely due to prior viral exposure. Vaccines that have received emergency use authorization in the US were designed to elicit an antibody response only against the SARS-CoV-2 S protein and for this reason N proteins detected in a serological assay are indicative of prior exposure to SARS-CoV-2 regardless of vaccine status (22, 23). If only S1 protein is detected in a serological assay and no N antibodies are detected, it is consistent with the subject having vaccine-induced antibody response.

A total of 27 subjects, 19 for Moderna and 8 for Pfizer-BioNTech, showed an immune response indicating previous exposure to SARS-CoV-2. These subjects had no prior positive SARS-CoV-2 antigen test, nor had they been symptomatic. After the first vaccination dose, the S1 IgG response in these subjects was increased (S/CO ∼ 25) as compared to subjects with no prior exposure (S/CO ∼ 15). S1 IgG response in subjects with known prior infection were significantly higher (S/CO > 100). This margin appeared to widen after the second vaccine dose. No IgG response was observed against S2 and N protein. Although, S2 is part of the vaccine S protein construct, S2 appears to have low immunogenicity as has been mentioned previously (24, 25). Vaccinated subjects were therefore differentiated into the following categories: no prior infection, known prior infection, and prior asymptomatic exposure/infection.

The [CDC has recently updated their guidance document](https://www.cdc.gov/coronavirus/2019-ncov/lab/resources/antibody-tests-guidelines.html) recognizing that serology assays may be utilized, “to help differentiate natural infection from vaccination by utilizing tests that measure antibodies against different protein targets.” More specifically, a serology test that is IgG positive for either N or S with or without vaccination indicates prior infection that could have occurred before or after vaccination. A serology test that is IgG positive for only S1 and negative for other antigens indicates no prior infection with SARS-CoV-2 as all S1 directed antibody resulted from a COVID-19 vaccination.

In this study, we set out to understand the utility of the multiplexed Quidel Sofia 2 SARS-CoV-2 IgG Antibody FIA that measures IgG antibodies against the three primary SARS-CoV-2 antigens S1, S2 and N from a single sample in 15 minutes. We conclude that fingerstick whole blood samples performed as well as serum, plasma, and venous whole blood samples. This would provide a distinct advantage to physicians in near-patient settings by providing accurate rapid results for their patients. The Sofia 2 SARS-CoV-2 IgG FIA is intended for use as an aid in identifying individuals with an adaptive immune response to SARS-CoV-2 S1, S2 and N proteins. Additionally, as this assay measures an IgG response against three viral proteins, S1, S2 and N, we can characterize immune response between *i)* naturally infected subjects, *ii*) vaccinated subjects with no prior infection, *iii*) vaccinated subjects with known prior infection, and *iv*) vaccinated subjects with prior asymptomatic exposure/infection. This study will provide invaluable insights into potential long-term protective humoral response as an increasing number of the world population is vaccinated against SARS-CoV-2.

## Methods

### Sofia 2 SARS-CoV-2 IgG Antibody FIA assay

The Sofia 2 SARS-CoV-2 IgG Antibody FIA is a single-use lateral flow rapid assay for the semi-quantitative detection of IgG antibodies to SARS-CoV-2 S1, S2 and N proteins. Sample types tested included fingerstick and venous whole blood, plasma, and serum derived from subjects prior to vaccination and at multiple time points after the first and second vaccinations. This assay utilizes three recombinant SARS-CoV-2 proteins that are separately immobilized on the lateral flow test strip. The recombinant N, S1 and S2 proteins consist of 425 amino acids, 681 amino acids and 539 amino acids, respectively. To perform the test with fingerstick and venous whole blood, a 25μL sample was collected into a disposable capillary device. This whole blood sample was diluted 40-fold and 100µl of this diluted sample was applied to the sample port of the test cassette containing the three immobilized antigens. For serum and plasma samples, 10μL of the sample was diluted 100-fold and 100μL was applied to the sample port of the test cassette. The SARS-CoV-2 IgG antibodies present in the sample, bind to fluorescent beads containing anti-human IgG antibodies and subsequently bind to the immobilized S1, S2 and N antigens. The test cassette was placed in the [Sofia 2 instrument](https://www.quidel.com/immunoassays/sofia-tests-kits/sofia-2-analyzer) and the IgG levels for each antigen were obtained at 15 minutes. The positive or negative values for each antigen are based on signal to cutoff ratios (S/CO) where S/CO ≥1 is positive and S/CO < 1 is considered negative.

### Statistical analysis

Data was normalized to the second vaccination day (Figure 2). LOESS smoothing with a span of 0.75 and degree of 2 was used to fit the longitudinal data of subjects within a category (21). R version 4.03 and RStudio version 1.4.1106 were used for study data analysis. Figures were generated using package ggplot2, version 3.3.3.

## Data Availability

This study is not part of a clinical trial.

## Acknowledgements

This study was funded by Quidel Corporation. We thank the contributions of the following Quidel employees: Adriana Gamboa, Nerliza Jimenez, Andrew Westberg, Bridget Balzon, Gabrielle Cruz, Allen Lalaian, Kenny Hwang, Lisa Pysz, Arianna Paik, Veronica Aziz, Kelly Brooks, Aaron Weston, Amanda Li, Cristina Charles, Minnie Chan, and Alexander Liu.

***QUIDEL*** *and* ***SOFIA*** *are registered trademarks of Quidel Corp. All other trademarks belong to their respective owners and their use herein does not imply sponsorship or endorsement of their products or services*.

